# Psychological state, family functioning and coping strategies among students of the University of Ibadan, Nigeria, during the COVID -19 lockdown

**DOI:** 10.1101/2020.07.09.20149997

**Authors:** Lucia Y. Ojewale

## Abstract

**Introduction:** The curtailment of social gatherings as well as lack of online academic engagement, during the COVID-19 lockdown, could have potentially damaging effects on the psychological state of university students in Nigerian public universities. This study examined the prevalence of anxiety and depression, including associated factors and coping methods among undergraduate students.

**Methods:** The study design was cross sectional and involved 386 undergraduate students. The study was approved with approval number UI/EC/20/0242. An online questionnaire was circulated among the students consisting mainly of Hospital Anxiety and Depression scale and the McMaster Family Assessment Device. Analysis was done using descriptive statistics, chi-square, Analysis of Variance (ANOVA) and linear logistics regression, at α 0.05.

**Results:** Mean age was 21±2.9 years, with a female population of 60.1%. Prevalence of anxiety and depression were 41.5% and 31.9% respectively. Students in health-related faculties were significantly less anxious than others. Inability to afford three square meals, negative family-functioning, having a chronic illness and living in a State/Region with high incidence of COVID-19, was significantly associated with depression. These factors jointly accounted for 14% of depression. Coping methods included use of social media, watching movies and participating in online skill development programmes.

**Conclusion:** There was a higher prevalence of anxiety and depression among university students with poor family functioning, inability to afford three meals/day, living in a state with a high incidence of COVID-19 and having a chronic illness. Proactive measures ought to be taken to support undergraduate students to prevent negative consequences of poor mental health.

## 1.0 Introduction

The novel coronavirus disease (COVID -19) pandemic, which was first detected in December 2019 has continued to ravage the entire world. The outbreak started in Wuhan, China[1] has since spread to almost all the countries of the world, [2]. Six months since the first case was reported, the number of confirmed cases has risen to over nine million - 9,633,157, [2]. Mortality from the disease is high and was estimated at 490,481 by June 27, 2020, [2]. Although the number of cases was slow to rise in Africa, they have been increasing exponentially, [3] in the last two months, threatening the already weak health structure, [4].

On March 11, 2020, the World Health Organisation (WHO) declared the novel Coronavirus a pandemic disease due to its spread all over the globe and how it has affected lifestyle and social interactions, [5]. Even though it started in China, it has rapidly spread throughout the world, affecting almost all countries of the world except isolated islands with few populations. It has been termed as a Public Health Emergency of International Concern by the WHO, [6].

The first case of coronavirus in Nigeria was confirmed on February 27 2020, [7], following which the Nigeria Centre for Disease Control (NCDC) issued several guidelines for curtailing with the spread, in line with the internationally-approved standard, [8]. These measures notwithstanding, the number of cases continued to rise and was estimated at 24, 077 by June 27, 2020, [9]. Already, by March 2020, the spread of the virus had led the federal and state governments and their various parastatals to shut down activities in the country. By March 23, both land border and air space had been closed; the weekly meeting of Federal executive council suspended, among other measures, [10]. On Monday, March 23 2020, the Nigerian University Commission (NUC), ordered the lockdown of all universities and affiliated schools, [11]. The lockdown has led to a halt in academic activities and students going home or discontinuing their industrial training - an unplanned break.

In addition to that, there was a lockdown of activities in three major states in the country from March 30, 2020, and a partial lockdown in most other states involving the cessation of all social and religious gatherings, [12]. The result was that Residents of Lagos and Ogun States and the Federal Capital Territory, in particular, where there was total lockdown, as well as those of other States, where there was partial lockdown had to spend most of their time at home.

Many employers have since made it possible for their employees to work from home while many private and secondary schools are taking measures to engage their students. Unfortunately, the reverse is the case among public tertiary institution students, who form the main bulk of Nigerian youths. Even before academic activities were halted due to the coronavirus pandemic, university staff across the country had already embarked on industrial action from March 9, 2020, [13]. Hence, there was no plan in place to engage public tertiary institution students, during the covid-19 lockdown, besides the inability to have industrial training/internship in companies due to the lockdown.

A feeling of being stalled in academic progress could lead to the development of anxiety or other psychological disorders since the period of youth is perceived as a period of opportunities, [14]. It is also not clear what role their family functioning may be playing in their psychological state. Given the increase in the incidence of mental disorders, particularly anxiety and depression, among young people, [15], examining the psychological state and coping mechanisms of undergraduate students in Nigeria has become crucial. This assertion is in line with a significant function of nurses in caring for the biopsychosocial state of individuals, whether sick or well.

Further, the WHO had reported in 2019 that mental health problems account for 16% of the global health burden among people aged 10-19 years, [16]. It further asserted that if left undiagnosed and untreated, it could lead to a myriad of physical and mental health issues hence preventing such individuals from living fulfilling lives, as adults, [16]. Some authors have reported an increased prevalence of anxiety and depression among university/college students in China and Greece during the lockdown, [17]–[20]. However, there is a dearth of information on the prevalence of anxiety and depression among undergraduate students in Nigeria during the COVID-19 lock down, as well as the associated. This study, therefore, reports the psychological effects of the lockdown, associated factors, family functioning and coping strategies among undergraduate students of the University of Ibadan, in Nigeria.

## 2.0 Materials and methods

### 2.1. Study design, setting and population

The descriptive study design was used, and the study was conducted among undergraduate students of the University of Ibadan, studying under the regular mode. The University of Ibadan is the first university in Nigeria and was founded on November 17 1948. It is located in Oyo State, south-western Nigeria. The institution, founded and funded mainly by the federal government, consists of sixteen faculties, [21]. Participants were drawn from all the faculties, which were merged into seven major faculties namely: Social Sciences; Clinical Sciences & Public Health; Pharmacy & Basic Medical Sciences; Law & Arts; Sciences & Technology; Education; Agriculture & Veterinary medicine.

### 2.2. Sample size and sampling technique

At a confidence level of 95% and a 4.94 confidence interval, 386 was obtained as a representative sample of the total undergraduate population of 20,000 using an automatic sample size calculator, [22]. A convenience sampling technique was utilised in getting in touch with participants via Whatsapp messaging platform.

### 2.3. Instrument

The study instrument consisted of a questionnaire made up of the following parts: Sociodemographic characteristics/ history of chronic illness, Hospital Anxiety Depression Scale (HADS), Family functioning Device and coping strategies. Questions on Sociodemographic characteristics included age, year of study, state of residence, whether there was a lockdown in the state of residence, among others. The State of residence were categorised into low, medium and high risk states based on the number of cases reported by the NCDC at the time of data collection. Moreover, as a result of the closure of many businesses due to measures to curtail the spread of the corona virus leading to a possible reduction in family income, participants were asked if they could afford three meals a day. Also, with regards to average weekly personal income, five options were provided: less than 1000; 1000 – 5000; 6000 – 10,000; 11,000 – 15 000; above 15,000 and these were reduced to two categories as most of the students chose the first option. These last two questions together with questions on ‘no of cars owned by the family’ and if the family were staying in their personal house were used to roughly estimate the socio-economic status of the students. The four questions, however, did not constitute a composite unit but were assessed individually. In order to determine the presence of chronic illness (s), students were asked if they had any of the following illnesses: asthma, sickle cell disease, diabetes, hypertension, kidney disease, depression and that they could tick more than one option.

The Hospital Anxiety Depression Scale (HADS) is a questionnaire made up of 14 items consisting of questions which evaluate anxiety and depression. It has been found suitable for use among psychiatric, non-psychiatric and healthy population, [23]. The authors also reported specificity and sensitivity of 0.8 for both sections of the questionnaire and the subscale scores range from 0-7 which denotes ‘normal’ or no anxiety/depression; 8-10 denoting ‘suspicious’ or mild and 11-21 pointing to the presence of anxiety or depression. These categories further showed that the higher the score, the more anxiety or depression is present. Moreover, the specificity and sensitivity of the scale have been established among Nigerians, [24], and this varied between 85% and 92.9%. The author, who carried out the study in both hospital and community, recommended the instrument for use in community surveys in developing countries. Another author had classified the scale scores into four, [25] but there was no data on the specificity and sensitivity based on these classifications, hence the three categories originally intended were used.

Family functioning Device (FAD): This is based on the McMaster Family Functioning model. The FAD is a scale whereby the structural and organisational characteristics of families are checked as well as the methods of interactions members of the family. It is suitable for differentiating between healthy and unhealthy families which could point to the availability or non-availability of social support. It has seven [7] subscales of Problem Solving, Communication, Roles, Affective Responsiveness, Affective Involvement, Behaviour Control and General Functioning scale. The scale is made available for free. The reliability α ranges from .72 to .83 for the subscales, and general functioning is .92. In this study, only the general functioning section consisting of 12 items was used. Responses are on a four-point Likert scale: “strongly agree,” “agree,” “disagree,” and “strongly disagree. Negatively-structured questions are reversed, and higher scores indicate problematic functioning. The highest obtainable score is 48, while the lowest obtainable score is 12. The higher the score, the poorer the family functioning, [26]

### 2.4. Data collection and analysis

After obtaining ethical approval from the University of Ibadan/University College Hospital ethical review board, the representatives of unions in the faculties were contacted through the students’ union vice president, who is a student at the Researchers’ department. The faculty executives then helped in getting across to their colleagues via online messaging platform, (Whatsapp). Subgroups within the faculties, including departments and levels, were also contacted through the faculty representatives. The data collected were transferred from the goggle form excel to Statistical Package for Social Sciences (SPSS) version 20.0 for analysis. Frequencies, percentages and mean were used in presenting the sociodemographic variables. These variables were tested for their association with anxiety and depression using the chi-square test and ANOVA. Those that were significant were entered into a linear regression model to determine to what degree they predict depression. Depression and anxiety were classified into three 0-7 which denotes ‘normal’ or absence of anxiety/depression; 8-10 denoting ‘suspicious’ or mild anxiety/depression and 11-21 pointing to the presence of moderate to severe anxiety or depression. These two measures were also tested for their association with family functioning using ANOVA. Post-Hoc analysis was carried out using Fisher’s Least Significant Difference to determine the pair means that were statistically different. Coping methods used by the students and their willingness to be seen by a health care provider if they were experiencing anxiety or depression were presented using bar charts.

### 2.5. Ethical considerations

The study protocol was submitted to the University of Ibadan/ University College Hospital Ibadan (U.I. /UCH) ethical review committee for ethical approval. The committee approved with approval number (UI/EC/20/0242). Besides, the questionnaire, circulated as Google form, included an introduction where the aims and benefits of the study were explained to the participants. Those who decided to complete the form gave consent implicitly.

## 3.0 Results

### 3.1. Sociodemographic characteristics and their association with anxiety and depression

Respondents’ sociodemographic characteristics and their association with anxiety and depression using the Chi-square test are presented in Table 1. The mean age of the undergraduate students was 20.8 (**±** 2.9) years, with a little over half (50.2%) in the 20-23 years age range. Most were female (60.1%), and Christianity was the most prevalent religion. Almost 20% of the participants could not afford three square meals. Eight percent (8%) of the participants suffered from one form of chronic illness or another, and 35.2% reported having a total lockdown in their state at the time the data was collected. Inability to afford three square meals and having a complete lockdown in the State of Residence was associated with depression, (p<0.001 and p<0.05, respectively). There was a significant association between students’ academic faculty and anxiety.

**Table 1:**
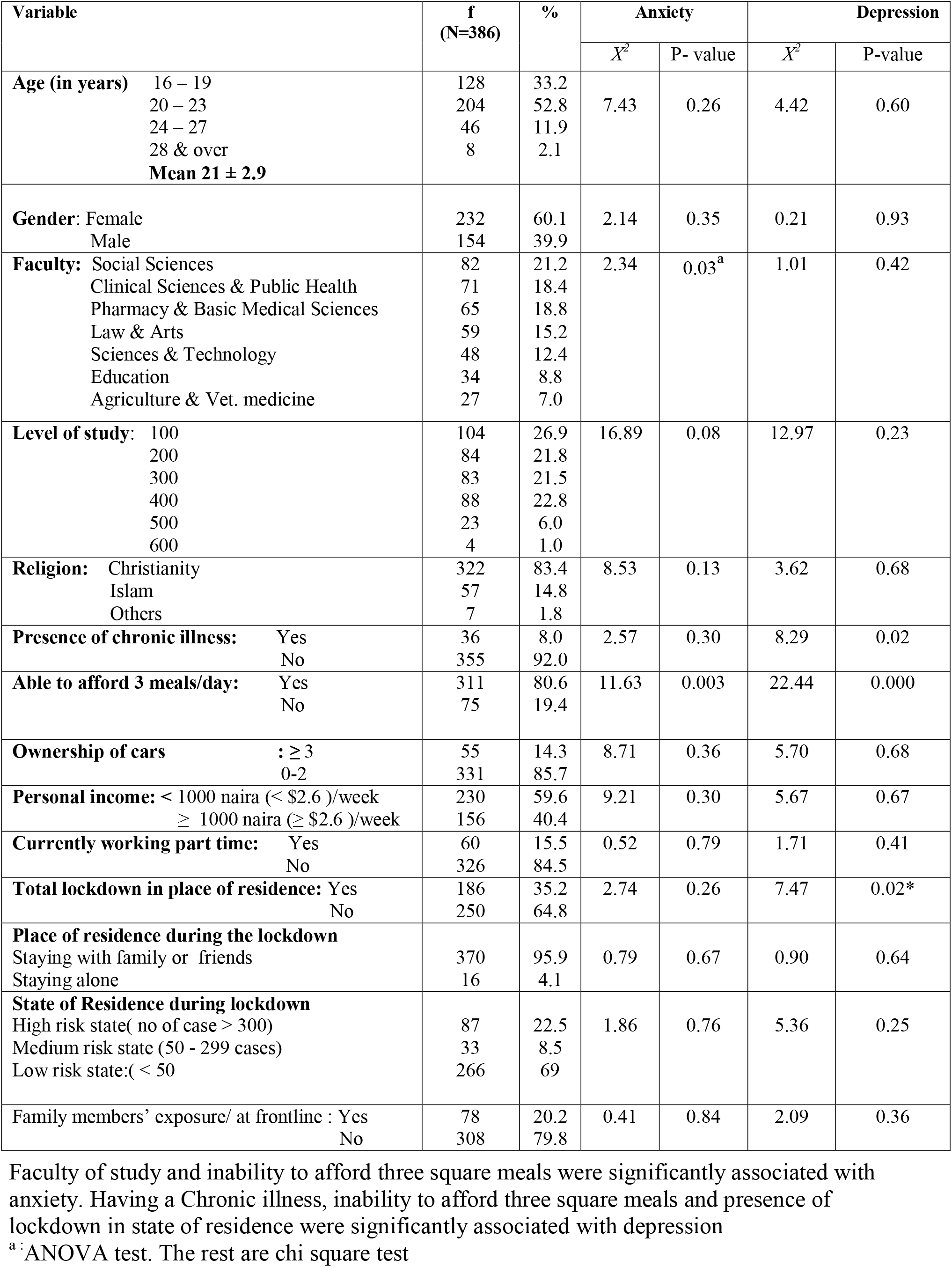
Analysis of association between Respondents’ sociodemographic characteristics and psychological state.

| Variable | f<br>(N=386) | % | Anxiety |  | Depression |  |
| --- | --- | --- | --- | --- | --- | --- |
|  |  |  | X<sup>2</sup> | P- value | X<sup>2</sup> | P-value |
| **Age (in years)** 16 – 19<br>20 – 23<br>24 – 27<br>28 & over<br>**Mean 21 ± 2.9** | 128<br>204<br>46<br>8 | 33.2<br>52.8<br>11.9<br>2.1 | 7.43 | 0.26 | 4.42 | 0.60 |
| **Gender:** Female<br>Male | 232<br>154 | 60.1<br>39.9 | 2.14 | 0.35 | 0.21 | 0.93 |
| **Faculty:** Social Sciences<br>Clinical Sciences & Public Health<br>Pharmacy & Basic Medical Sciences<br>Law & Arts<br>Sciences & Technology<br>Education<br>Agriculture & Vet. medicine | 82<br>71<br>65<br>59<br>48<br>34<br>27 | 21.2<br>18.4<br>18.8<br>15.2<br>12.4<br>8.8<br>7.0 | 2.34 | 0.03<sup>a</sup> | 1.01 | 0.42 |
| **Level of study:** 100<br>200<br>300<br>400<br>500<br>600 | 104<br>84<br>83<br>88<br>23<br>4 | 26.9<br>21.8<br>21.5<br>22.8<br>6.0<br>1.0 | 16.89 | 0.08 | 12.97 | 0.23 |
| **Religion:** Christianity<br>Islam<br>Others | 322<br>57<br>7 | 83.4<br>14.8<br>1.8 | 8.53 | 0.13 | 3.62 | 0.68 |
| **Presence of chronic illness:** Yes<br>No | 36<br>355 | 8.0<br>92.0 | 2.57 | 0.30 | 8.29 | 0.02 |
| **Able to afford 3 meals/day:** Yes<br>No | 311<br>75 | 80.6<br>19.4 | 11.63 | 0.003 | 22.44 | 0.000 |
| **Ownership of cars** : ≥ 3<br>0-2 | 55<br>331 | 14.3<br>85.7 | 8.71 | 0.36 | 5.70 | 0.68 |
| **Personal income:** < 1000 naira (< \$2.6 )/week<br>≥ 1000 naira (≥ \$2.6 )/week | 230<br>156 | 59.6<br>40.4 | 9.21 | 0.30 | 5.67 | 0.67 |
| **Currently working part time:** Yes<br>No | 60<br>326 | 15.5<br>84.5 | 0.52 | 0.79 | 1.71 | 0.41 |
| **Total lockdown in place of residence:** Yes<br>No | 186<br>250 | 35.2<br>64.8 | 2.74 | 0.26 | 7.47 | 0.02* |
| **Place of residence during the lockdown**<br>Staying with family or friends<br>Staying alone | 370<br>16 | 95.9<br>4.1 | 0.79 | 0.67 | 0.90 | 0.64 |
| **State of Residence during lockdown**<br>High risk state( no of case > 300)<br>Medium risk state (50 - 299 cases)<br>Low risk state:( < 50 | 87<br>33<br>266 | 22.5<br>8.5<br>69 | 1.86 | 0.76 | 5.36 | 0.25 |
| **Family members' exposure/ at frontline :** Yes<br>No | 78<br>308 | 20.2<br>79.8 | 0.41 | 0.84 | 2.09 | 0.36 |

Faculty of study and inability to afford three square meals were significantly associated with anxiety. Having a Chronic illness, inability to afford three square meals and presence of lockdown in state of residence were significantly associated with depression
<sup>a</sup>: ANOVA test. The rest are chi square test

Post Hoc analysis [LSD – Least Significant Difference (LSD)] of mean scores among the different faculties showed that students in the faculties of Clinical sciences/Dentistry/Public health showed a significantly lower level of anxiety (5.2 ±4.1) compared to their counterparts in Social sciences and economics (6.61±4.50, p = 0.046); Science and Technology (7.4±4.5, p = 0.01); Education (7.7 ± 4.1, p = 0.01) and Agriculture/veterinary medicine 7.9 ±5.4, p = 0.01). However, there was no significant difference between the scores of those in Clinical sciences/Dentistry/Public health and Pharmacy/Basic Medical Sciences (5.7±3.4), p = 0.64

Additionally, students in the faculty of education showed a higher mean score on depression (7.2±3.9) compared to students in the faculties of Pharmacy/Basic Medical Sciences (5.7±3.4), p= 0.04.

### 3.2. Prevalence of anxiety and depression and willingness to consult a health professional

Table 2 shows that 20.7% of the students had moderate to severe anxiety, with another 20.7% suffering from mild anxiety. Also, the prevalence of moderate to severe anxiety was 10.9%, while 21% had mild depression.

**Table 2:**
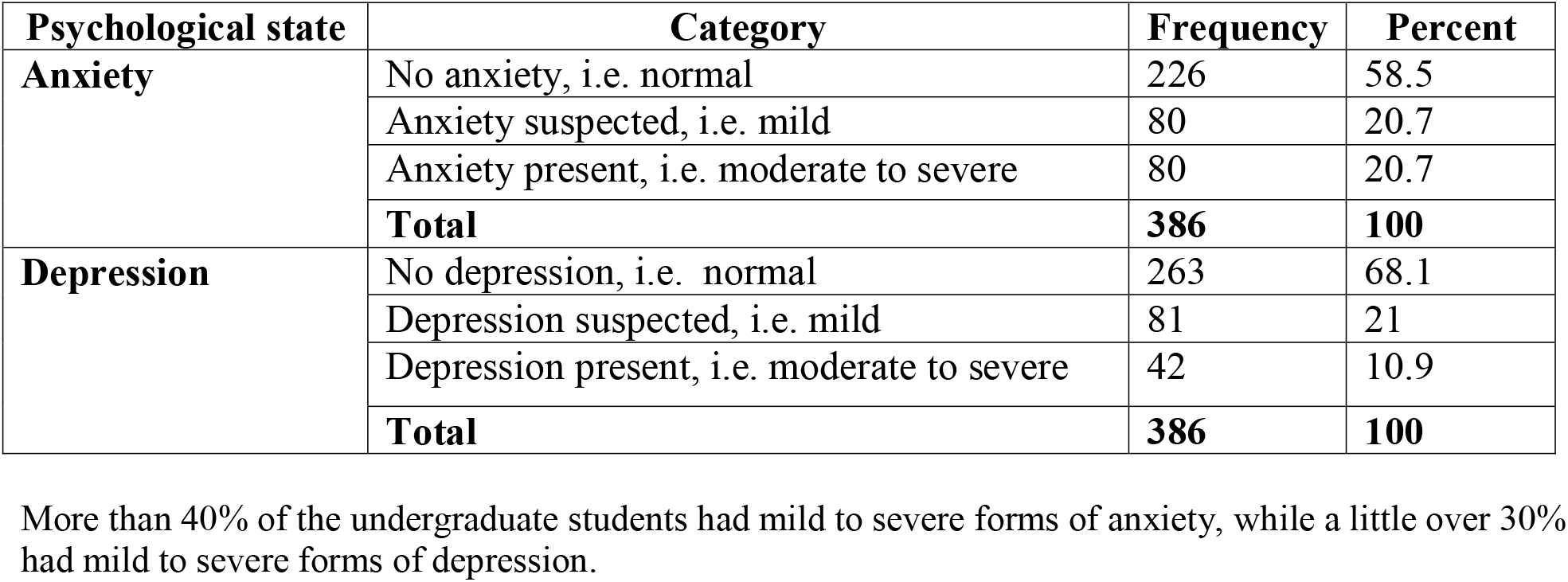
Prevalence of anxiety and depression.

| Psychological state | Category | Frequency | Percent |
| --- | --- | --- | --- |
| **Anxiety** | No anxiety, i.e. normal | 226 | 58.5 |
|  | Anxiety suspected, i.e. mild | 80 | 20.7 |
|  | Anxiety present, i.e. moderate to severe | 80 | 20.7 |
|  | **Total** | **386** | **100** |
| **Depression** | No depression, i.e. normal | 263 | 68.1 |
|  | Depression suspected, i.e. mild | 81 | 21 |
|  | Depression present, i.e. moderate to severe | 42 | 10.9 |
|  | **Total** | **386** | **100** |

More than 40% of the undergraduate students had mild to severe forms of anxiety, while a little over 30% had mild to severe forms of depression.

Fig 1 shows that about half of the students (48.4%) were willing to see a health care professional if they were feeling anxious or depressed.

**Fig 1:**
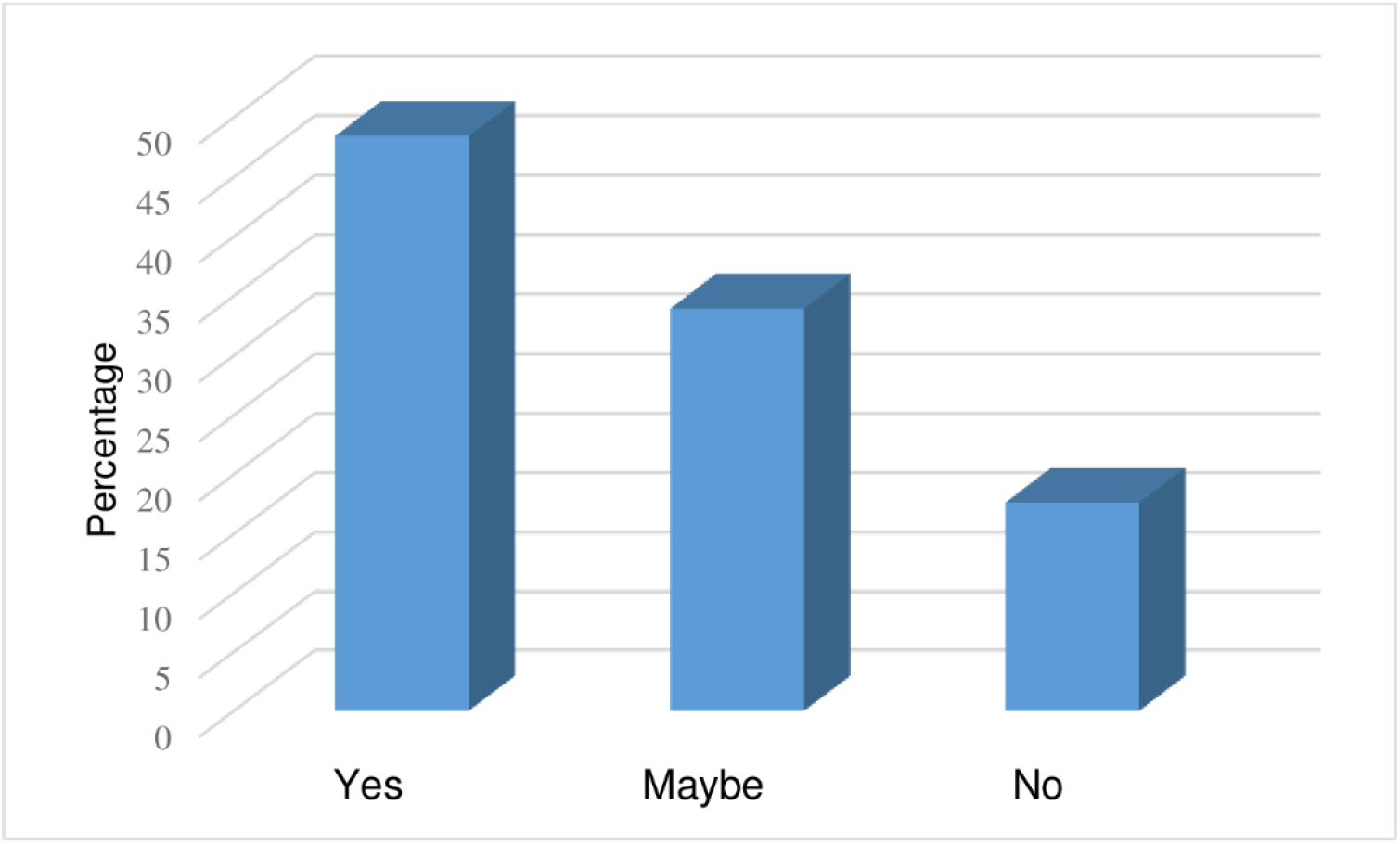
Students’ willingness to speak to a health care professional.

A good number of students were open to discussing their mental health state with a health professional

### 3.3. Coping with the lockdown

As shown in Fig 2, students coped with the lockdown and cessation of academic activities by being on social media (17.9%), watching television or videos (11.1%), engaging in online skill-building activities (15.3%). Other methods of coping included sports activities, meditation, helping at home, games and music.

**Fig 2:**
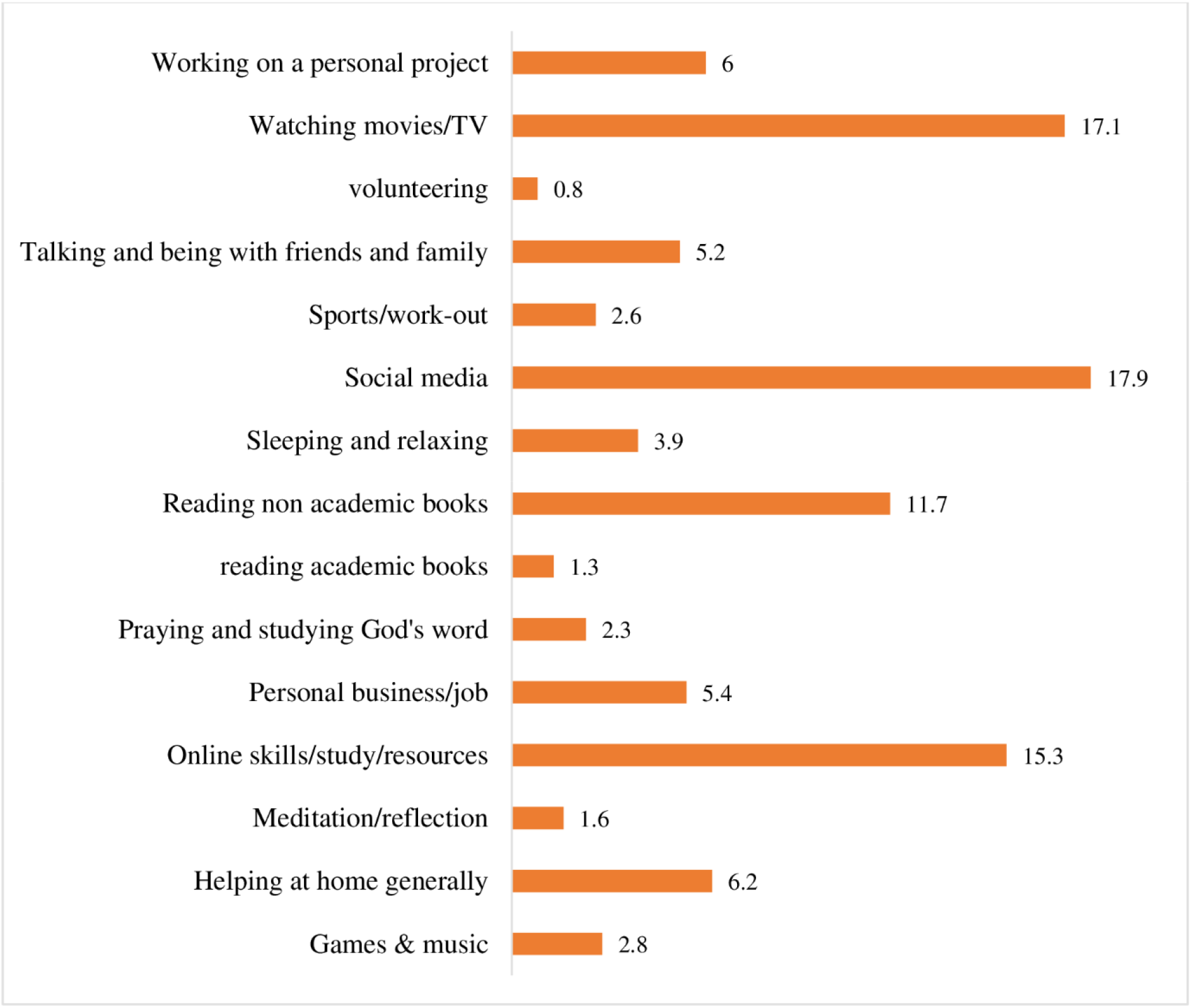
Students’ coping method.

Students’ best activity for coping with the lockdown included virtual entertainment, skill acquisition and meditation

### 3.4. Association between family functioning and psychological states

Table 3 shows that there was a significant effect of family functioning of students on presence, suspected presence or absence of anxiety at p < 0.01 [F (2, 383) = 15.49, p = 0.000], using Analysis of Variance (ANOVA)

**Table 3:** ANOVA showing association between family functioning and anxiety.

|  | **N** | **Mean** | **Std. Deviation** | **F** | **P - value** |
| --- | --- | --- | --- | --- | --- |
| No anxiety | 226 | 23.2 | 5.1 | 15.49 | 0.000<sup>a</sup> |
| Anxiety suspected | 80 | 24.8 | 6.0 |  |  |
| Anxiety present | 80 | 27.2 | 6.5 |  |  |

The mean difference between undergraduate students who were anxious or had suspected anxiety was significantly different.
<sup>a</sup> significant at $P < 0.01$ .; two-tailed

Moreover, the Post-Hoc comparison of the mean scores of the three categories, using Fisher’s Least Significant Difference (LSD), (Table 4), revealed that the mean score for family functioning among students with no anxiety (23.2 ± 5.1) was significantly different from that of those in whom anxiety was suspected (24.8 ± 6.0); p = 0.03. Similarly, there was a significant difference between those with no anxiety (23.2± 5.1) and those in whom anxiety was present (27.2 ± 6.3, p = 0.000). Higher scores denote poor family functioning.

**Table 4:** Post Hoc Analysis for family functioning and anxiety state.

| **Anxiety Category** | **Anxiety category** | **Mean Difference** | **P-value** | **95% Confidence Interval** |  |
| --- | --- | --- | --- | --- | --- |
|  |  |  |  | **Lower Bound** | **Upper Bound** |
| No anxiety | Anxiety suspected | -1.564 | 0.03 | -2.99 | -0.13 |
|  | Anxiety present | -4.014 | 0.000<sup>a</sup> | -5.44 | -2.58 |
| Anxiety suspected | No anxiety | 1.564 | 0.03 | 0.13 | 2.99 |
|  | Anxiety present | -2.450 | 0.01 | -4.19 | -0.71 |
| Anxiety present | No anxiety | 4.014 | 0.000<sup>a</sup> | 2.58 | 5.44 |
|  | Anxiety suspected | 2.450 | 0.01 | 0.71 | 4.19 |

The family functioning of students who were not anxious at all, was significantly different from that of students who were suspected of having anxiety at $p < 0.05$ and from those with anxiety at $p < 0.01$ . There was also a significant difference in the family functioning of students in whom anxiety was suspected and those with actual anxiety.
<sup>a</sup> $p < 0.01$

ANOVA results on depression and family functioning (Table 5) showed a significant association between the two variables [F (2, 383) = 14.209, p = 0.000].

**Table 5:** ANOVA showing association between depression and family functioning.

| Category | N | Mean | Std. dev. | F | P-value |
| --- | --- | --- | --- | --- | --- |
| No depression | 263 | 23.4 | 5.4 | 14.209 | 0.000<sup>a</sup> |
| Depression suspected | 81 | 25.9 | 5.9 |  |  |
| Depressed | 42 | 27.6 | 6.1 |  |  |

There was a significant association between family functioning and depression.
<sup>a</sup> significant at $p < 0.01$ ; two-tailed.

Besides, the Fisher’s Least Significant Difference (LSD), in Table 6 showed that the mean score for family functioning among students without depression (23.35 ± 5.44) was significantly different from that of students who had borderline (suspected) depression (25.88 ± 5.87); p = 0.000; as well as those who were depressed (27.6 ± 6.13), p = 0.000.

**Table 6:** Post Hoc Analysis for family functioning and depression.

| Depression category | Depression category | Mean Difference | P - value | 95% Confidence Interval |  |
| --- | --- | --- | --- | --- | --- |
|  |  |  |  | Lower Bound | Upper Bound |
| No depression | Depression suspected | -2.527 | 0.000<sup>a</sup> | -3.93 | -1.13 |
|  | Depressed | -4.245 | 0.000<sup>a</sup> | -6.08 | -2.41 |
| Depression suspected | No depression | 2.527 | 0.000<sup>a</sup> | 1.13 | 3.93 |
|  | Depressed | -1.719 | 0.11 | -3.82 | 0.38 |
| Depression | No depression | 4.245 | 0.000<sup>a</sup> | 2.41 | 6.08 |
|  | Depression suspected | 1.719 | 0.11 | -0.38 | 3.82 |

Students who were depressed had a significantly better family functioning than students who had suspected and actual depression.
<sup>a</sup> $p < 0.01$

However, there was no significant difference between the family functioning of those with borderline depression (25.88 ± 5.87) and those who were depressed (27.6 ± 6.13), p = 0.108.

### 3.5. Multiple linear regression on variables that predict depression

A multiple linear regression was performed to determine to what extent some variables predict the presence of depression, as shown on Table 7. Results indicated that the model explained 14.3% of the variance and that the model was a significant predictor of depression, F (4,381) = 15.92, p < .001. The results also show that three meals/day, [Beta = 0.19, t (381) = 3.97, p < 0.01]; chronic illness [Beta = 0.123, t (381) = 2.5, p = 0.01] and family functioning [Beta = 0.264, t (381] = 5.52, p < 0.01] significantly predict presence of depression. However, having lock down in the state where the students were currently resident was not statistically significant.

**Table 7:** Multiple linear regression analysis on the relationship between selected variables and depression.

|  | **B** | **Std. Error** | **$\beta$** | **t** | **P -value** |
| --- | --- | --- | --- | --- | --- |
| (Constant) | -2.577 | 1.189 |  | -2.168 | 0.03 |
| 3 meals/day | 1.683 | 0.424 | 0.19 | 3.97 | 0.000 |
| lockdown in State | 0.614 | 0.35 | 0.084 | 1.752 | 0.08 |
| Chronic illness | 1.589 | 0.614 | 0.123 | 2.59 | 0.01 |
| Family functioning | 0.16 | 0.029 | 0.264 | 5.517 | 0.000 |

$F(4,381) = 15.92, p < .001, R^2 = 0.143 (14.3\%), R^2_{\text{Adjusted}} 0.134$
The model shows that the four factors could predict 14% of depression seen in the undergraduate students.

Participants’ predicted depression score is equal to - 2.58 + 1.68(3 meals/day) + 0.61 (lockdown in state of residence) + 1.59 (chronic illness) + 0.16 (family functioning); where 3 meals/day and lockdown in state of residence are codes as 2 = Yes, 1 = No; and chronic illness is coded as 2 = Yes, 1 = No. Students who were unable to afford three square meals, who had a chronic illness (s) and who had a poor family functioning were more depression.

## 4.0. Discussion

The lockdown associated with the coronavirus pandemic is an exceptional event in Nigeria. Every sector of the country has been affected, although the Federal and State Governments were gradually easing the lockdown. As of the end of June 2020, when this report was being written, the undergraduate students had not had any form of academic engagement, for almost three months.

The response rate for the survey was high as the sample size of 386 was reached within a week. Most of the participants were female, (60%), although the male: female student ratio in the school is usually around 53:47, [27]. The higher percentage of females who participated in the study could be associated with the higher interest of women in psychology-related issues. The dominance of women in psychology is supported by Clay [28], who stated that there are more females in the field of psychology – both in education and the workforce. Similarly, Cope et al.,[29] found a significantly higher number of females enrolled for postgraduate programmes in psychology than male, as high as 75% female compared to 25% male; a trend which had been occurring for about ten (10) years.

Students with one or more chronic illness (s) had a significantly higher level of anxiety and depression compared with those who did not have such a disease. This is supported by Wang et al., [30], who reported a higher level of psychological distress among people with a medical history of chronic illness in a population-based study involving 1210 people in China.

Contrary to the findings of some authors, [18, 19, 31], in which female participants had a significantly higher level of depression than their male counterparts, during the covid-19 pandemic; there was no significant association between gender and anxiety or depression among the participants in this study. The difference may be associated with the variation in the demography. Whereas, the present study was carried out among university students aged 16 to 30 years; these previous authors, [18, 19, 31] carried out their survey among all age groups. The present findings are in keeping with that of some other authors, [17, 32, 33], which were all carried out among university students. It is possible that other factors, such as age, combine to make females more prone to psychological distress. However, another study, [34] among university students in China showed a significantly higher prevalence of depression among female students.

Although measures of socioeconomic status including the number of cars owned by the family; private income and living in a personal house were not associated with anxiety and depression; students who could not afford three square meals/day had a significantly higher level of anxiety and depression. Often though, being able to provide three meals is linked to income. Hence it can be presumed that students who could not afford three square meals were of low socioeconomic background. Cao et al. [17] had also reported that poor socioeconomic status was associated with anxiety among the university students who participated in their study, with those of poor economic conditions experiencing more considerable anxiety. At the beginning of the lockdown, the federal government and some state governments had promised palliative measures to citizens of the country to ease the economic burden of the lockdown. However, due to limited resources, the palliative measures could only be provided to the ‘poorest of the poor’. This socioeconomic group is mostly resident in the rural parts of the country and constitute 49.66% of the population, [35]. Therefore, those in the urban areas, which is where most university students live, were not provided with palliatives, hence the inability to have three square meals.

On the prevalence of anxiety, whereas Cao et al. [17] reported that about 25% of Chinese university students had mild to severe forms of anxiety during the COVID -19; in this study, a higher percentage of Nigerian university students (41.5%) experienced mild to severe forms of anxiety. The higher rate could be associated with different periods of data collection. While Cao et al. [17) collected data in February 2020, before the pandemic reached its peak in most parts of the world, data collection for this study occurred in May 2020, a period during which the cases were increasing exponentially in Nigeria. Some authors, [36], had reported a lower rate of anxiety, 14.8%, before the COVD-19 pandemic, among undergraduate students in five universities in South-western Nigeria. Again, this is not surprising since several studies that took place in Italy, United States, China and Spain, [18–20, 32, 37], have averred that the lockdown/self-quarantine associated with the COVID-19 was putting a lot of psychological strain on college students

Further, mild to severe depression was present in 31.9% of university students who participated in this study, of which one-third were adolescents (aged 16-19 years). There was no significant association between the age categories and depression. Therefore this prevalence could be applied to the adolescent age group who participated in this study. Although, there is no national data on the prevalence of depression among Nigerian youths, there are reports of studies carried out in the same geopolitical zone where the present study was conducted. For instance, the prevalence found in this study is higher than that earlier reported for Nigerian adolescents, [38],in which 21.2% of rural adolescents in south-western Nigeria, were found to have varying degrees of depression. It is not surprising that more participants in this study experienced depression since the COVID-19 lockdown was something that put emotional strain on many people. Likewise, a somewhat lower prevalence rate of depression – 21.6% - was reported by Ayandele et al. [36)] in 2019, among undergraduate students in five universities located in the same region where the present study took place.

Earlier on, in 2013, Peltzer et al. [39] had documented a prevalence of 25.2% of moderate to severe forms of depression among undergraduate students in Western Nigeria. However, 10.9% of undergraduate students who participated in this study experienced moderate to severe depression. This variation could be because participants in the study of Peltzer et al. [39] experienced more depression as a result of other sensitive information such as the experience of sexual abuse, physical violence or injury, use of substances were elicited from the participants at the time data that was collected on depression. This information might have brought on some negative feelings leading to a higher score on depression.

Moreover, students who lived alone did not experience a significantly higher level of anxiety compared with those living with family or friends. Mazza et al. support this finding, [18]. Since most participants were young adult, it could be that those living alone were able to fend for themselves and therefore did not suffer from a significantly higher level of anxiety. Furthermore, results of Post-Hoc ANOVA, showed that students in non-health related faculties had a considerably higher level of anxiety than those in health-related fields. This interfaculty difference is congruent with what was reported in another study among college students in China and Spain [34, [37].

Another contributory factor to depression among undergraduate students was family functioning. Participants with poor family functioning experienced mild to severe depression, while those with good family functioning were not depressed. As mentioned earlier, staying alone or with family or friends did not significantly influence the experience of depression, yet staying with family or friends, as almost all the participants did, while experiencing poor functioning was a significant factor causing depression. Cao et al. [17] had reported a negative correlation between social support and anxiety, such that as social support increases, anxiety reduces. Further, Li et al. [40], reported that good family function was associated with a decreased level of psychological distress among health professional students. This was also the position of Jianjun, et al. in a study among undergraduate students in China [41]

Besides, the top three coping methods used by the undergraduate students included social media-Whatsapp, status challenges, Instagram, among others - watching television or movies and participation in online skills acquisition programmes or extra-curricular study activities. These methods are similar to those mostly used by university students in Jordan, as reported by Al-Tamemmi et al. [[33]. Also, students in states where there was a total lockdown experienced a significantly higher level of depression. This difference is logical since states with lockdown had a higher number of cases, and the students must have experienced increased susceptibility to the coronavirus infection coupled with lack of social gatherings or religious interaction. This is in consonant with what was reported by some authors, in studies involving higher institution students in China, [42,43]. Another interesting finding is the students’ responses regarding their ‘willingness to speak to a health professional if they were experiencing anxiety or depression. Almost half, (48.4%), of the students answered in the affirmative, while about a fifth (17.6%)answered ‘No’; the rest were undecided. Two studies carried out in 2017, and 2018 [44, [45] had shown a negative attitude towards mental illness among young people in a secondary school and a University in Nigeria, respectively. Another study reported a positive attitude by Pharmacy students [46].

On the whole, factors that were significant from the univariate analysis and included in the linear regression showed that that not being able to afford three meals in a day; poor family functioning; having a chronic illness and having a total lockdown in the state of residence accounted for 14% of depression seen among participants in the study.

## Conclusion

In conclusion, the lockdown associated with COVID-19 has placed a lot of psychological stress on university students as more than a third of those who participated in this study had varying degrees of anxiety and depression. It is recommended that measures such as financial palliative or provision of food to needy families, counselling of dysfunctional families and appropriate education and support of students with chronic illnesses, be put in place to alleviate the psychological distress of these students. There is also a need for recommencement of academic activities through online engagement to positively channel the energy of the university students.

## Data Availability

All data referred to in the manuscript are available in the link provided below

https://docs.google.com/forms/d/1_baBKFvrQTkxMd6AwXiSFw5TAY9OZhRGNnRQDyemXMA/edit#responses

## Study limitations

The Hospital Anxiety and Depression scale (HADs) used in data collection does not differentiate between moderate and severe forms of the two psychological states.

## Acknowledgement

The author specially thanks Professor G.O. Ademowo of the Institute of Institute for Advanced Medical Research and Training (IMRAT), College of Medicine, University of Ibadan for guidance regarding the online ethical application process. The author also acknowledges students’ faculty representatives who facilitated the data collection. These are Oluwatumininu Oma, Olamide Akinyemi, Temilola Akinditi, Chisom Okafor, Adanna Emeaghara, Elizabeth Okoye, Nkechi Nweya, Judith Onyekwere, Udochukwu Okeahialam, Chibueze Agwuncha, Ahmed Ajibade, Irewole Olumeyan, Adeola Olajide, Olalekan Michael Ajibola, John Fadele, Iyanuoluwa, Kene and Wole.

